# Shoulder muscle synergies before and after reverse total shoulder arthroplasty

**DOI:** 10.1101/2025.08.29.25334185

**Authors:** H. Francalanci, N. Holzer, F. Moissenet, Y. Cherni

**Affiliations:** Laboratoire de neurobiomécanique et neuroréadaptation de la locomotion, Centre de recherche Azrieli du CHU Sainte Justine, Montréal, Canada; Orthopedic Surgery and Musculoskeletal Trauma Care Division, Department of Surgery, Geneva University Hospitals, Geneva, Switzerland; Biomechanics Laboratory, Geneva University Hospitals and University of Geneva, Geneva, Switzerland; Kinesiology Laboratory, Geneva University Hospitals and University of Geneva, Geneva, Switzerland; Ecole de kinésiologie et sciences de l’activité physique, Université de Montréal, Montréal, Canada; Institut de génie biomédical, Université de Montréal, Montréal, Canada

**Keywords:** Reverse total shoulder arthroplasty, Massive rotator cuff tear, Motor control, Synergies, Electromyography

## Abstract

**Background:** Reverse total shoulder arthroplasty (rTSA) has become a standard treatment for symptomatic massive rotator cuff tears (MRCT). While clinical outcomes are generally satisfactory, significant variability in functional recovery raises questions about the extent of inter-individual neuromuscular adaptations and motor restoration. This study aimed to assess the neuromuscular impact of rTSA by analysing superficial shoulder muscle activity and synergies.

**Methods:** Asymptomatic shoulders (n=20) and symptomatic shoulders undergoing rTSA (n=20) were assessed before and after surgery. Participants performed four functional tasks while surface electromyography was used to quantified muscle activity. Synergies of the anterior (DELTA), middle (DELTM), and posterior deltoid (DELTP), middle (TRAPM) and upper trapezius (TRAPS), and serratus anterior (SERRA) were computed.

**Results:** Symptomatic MRCT was associated with a significant increase in activity of DELTM, TRAPS, and SERRA (p < 0.001). Postoperatively, DELTM (p = 0.003) and TRAPS (p = 0.039) remained significantly more active. Although the number of muscle synergies (n = 2) remained stable across groups, significant differences in muscle weightings were found preoperatively for TRAPS and DELTP (p < 0.001). Also, cosine similarity increased postoperatively by 15.96% for synergy 1 and 37.42% for synergy 2, indicating a greater resemblance to the asymptomatic synergy pattern (p < 0.001).

**Conclusion:** Changes in muscles activities and synergy patterns, marked by the emergence of compensatory roles for scapular stabilizers and the deltoid, highlight the disruption of motor control associated with MRCT. Following rTSA, motor coordination was substantially restored, supporting improved functional autonomy in daily tasks.

**Highlights:**

- Muscle activation and synergy analyses reveal altered motor control in shoulders with massive rotator cuff tear.
- Deltoid and scapular stabilizers show compensatory roles preoperatively.
- Reverse total shoulder arthroplasty leads to partial restoration of physiological coordination patterns.
- Synergy profiles shift significantly toward asymptomatic motor strategies after surgery.

## 1. Introduction

The motor system’s ability to reorganise muscle activation patterns is a key factor in restoring motor function in patients with shoulder disorders. This neuromuscular organisation relies on the interaction between ligamentous and muscular structures that ensure both mobility and stability to the glenohumeral joint [1,15]. A central component of this balance is the rotator cuff, a group of 4 muscles and related tendons responsible for the dynamic stabilisation of the shoulder, required for efficient joint mobilisation by the deltoid muscle [28]. When the cuff is torn, particularly in cases of massive rotator cuff tears (MRCT), motor control mechanisms become disrupted [16,47], leading to the loss of rotator cuff forces equilibrium, superior migration of the humeral head, and decrease of deltoid efficiency [21]. As a result, shoulder joint forces imbalance may lead to compensatory muscle activation strategies, which ultimately reduce functional efficiency and impair muscle coordination [11,12].

By restoring a stable centre of rotation of the glenohumeral joint, reverse total shoulder arthroplasty (rTSA) enables the deltoid to become the principal contributor of arm elevation in the absence of rotator cuff [21]. While this reconfiguration allows significant improvements in active elevation and functional independence [45,52], it also results in significant neuromuscular adaptations. An hyperactivation of the deltoid is frequently observed, often accompanied by a compensatory activity from adjacent muscles such as the upper trapezius, serratus anterior, and latissimus dorsi [27,39,46]. In addition to these neurophysiological changes, the considerable inter-individual variability raises concerns about the overall effectiveness of motor recovery following rTSA [27,34].

In this context, electromyography (EMG) is considered a standard for quantifying muscle activity and examining postoperative adaptations. However, beyond the magnitude of muscle activation, it is the pattern of coordinated muscle recruitment that offers deeper insight into the underlying motor strategy. The concept of muscle synergies, introduced by Bernstein [2], serves as a useful approach for modelling motor control. It suggests that the central nervous system, through mechanisms located in the spinal cord, activates groups of muscles simultaneously and in a coordinated manner during movement execution [10]. Analysing muscle synergies can thus help in identifying motor patterns and tracking their evolution across various clinical contexts [36]. In patients with musculoskeletal disorders of the upper limb, studies have shown that both the number and structure of these muscle synergies can be altered [14], reflecting either a motor control disruption or the implementation of compensatory strategies. However, to our knowledge, muscle synergy analysis has not yet been applied in the context of rTSA. When used in this population, it can offer the opportunity to understand how the motor system adapts to pathological state and therapeutic approaches. Specifically, it may allow for the assessment of whether postoperative muscle coordination patterns resemble those of asymptomatic shoulders or reveal altered compensatory strategies.

This study aims to analyse the evolution of shoulder muscle activity and synergies before and after rTSA, based on EMG data collected during standardised functional tasks. By comparing asymptomatic, preoperative, and postoperative shoulders, it proposes a neuromuscular perspective on motor recovery. The primary objective is to better characterise the muscle coordination strategies implemented after surgery, beyond the assessment of motor performance alone. We hypothesize that rTSA does not significantly alter the number or organisation of muscle synergies compared to asymptomatic shoulders, which would suggest a global motor control restoration.

## 2. Method

### 2.1. Participants

This retrospective study included EMG data from patients who underwent elective rTSA at Geneva University Hospitals between 2022 and 2025. All participants provided written informed consent prior to inclusion, in accordance with the Declaration of Helsinki. Ethical approval was obtained from the Geneva University Hospitals Ethics Review Board (CER 2025-00773).

Two shoulder groups were defined: (1) asymptomatic contralateral shoulders, with normal range of motion [51], no pain, and no history of musculoskeletal pathology or surgery; and (2) MRCT shoulders, assessed preoperatively and at 12 months postoperatively, with MRI-confirmed tears and treated with autologous bone grafting to correct glenoid orientation, targeting a baseplate neutral version and −5° inferior inclination. Exclusion criteria included revision surgeries, concomitant shoulder disorders, or inability to complete the protocol. Adequate EMG signal quality was required in both groups (see section 2.4.1). Because the number of synergies is considered a relatively insensitive variable, the sample size estimation was based on the variance accounted for by the first synergy (VAF1). Based on findings from Essers et al. [14] who reported a 13-point difference in VAF1 between symptomatic and asymptomatic groups (74% vs. 87%), a power analysis for a two-tailed paired t-test (α = 0.05, effect size = 0.65, power = 80%) indicated a minimum of 20 shoulders per group.

### 2.2. Procedure

Participants were seated in a standardised position on a chair and performed eight motor tasks, each repeated three times, with a 3-second rest period between repetitions (Fig. 1). These tasks were divided into two categories: (1) analytical tasks (only used for EMG signal normalisation), including bilateral flexion in the sagittal plane, abduction in the frontal plane, and both external and internal rotations at 0° abduction; and (2) functional tasks, including bring the hands to the mouth, to the top of the head, to the highest achievable point above the head, and to the highest possible point along the spine behind the back (or as close as possible if full range was not achievable). All experimental sessions were supervised by the same experienced operator (FM) to ensure consistency in the protocol execution and data collection.

**Figure 1:**
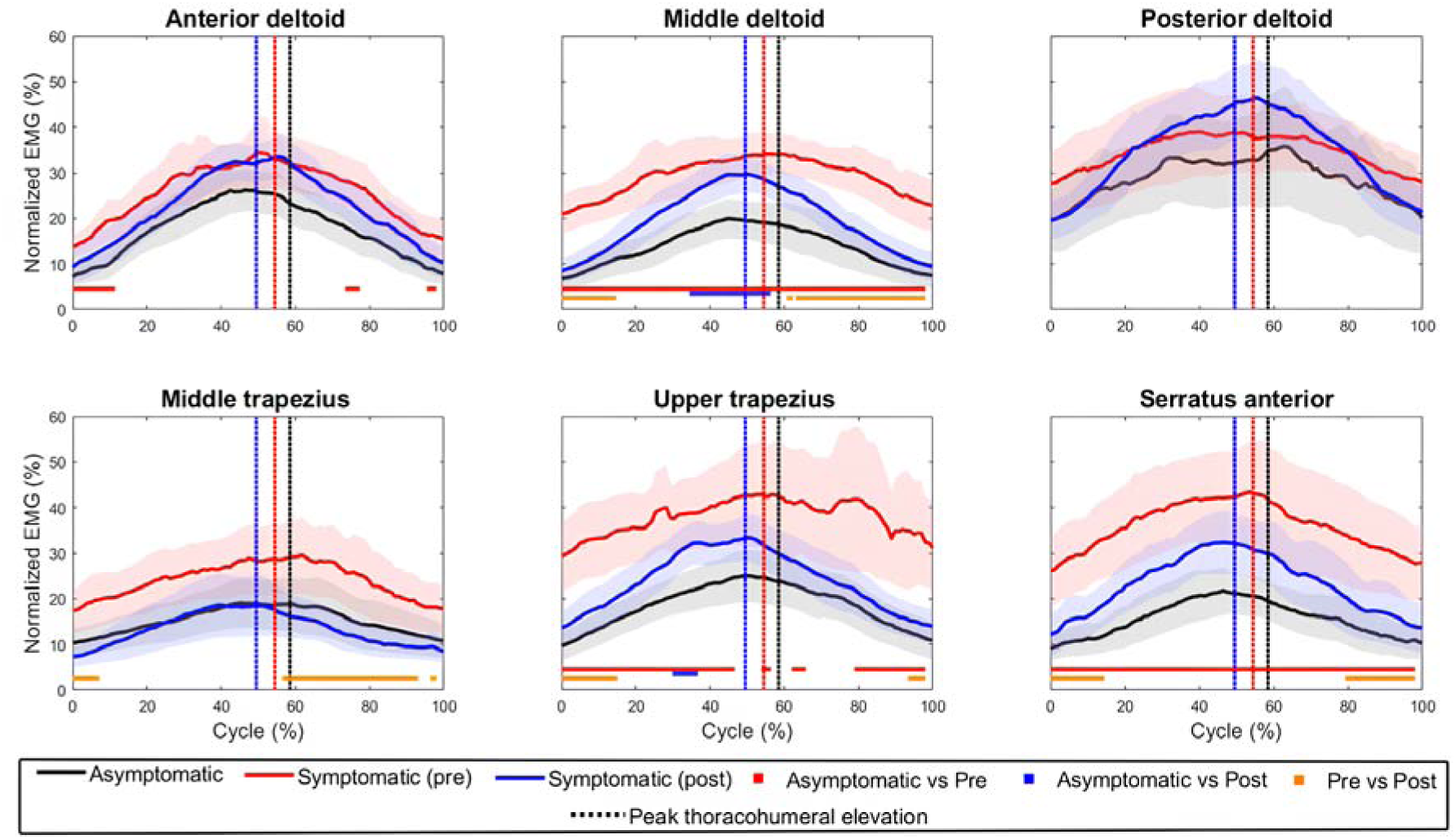
Intergroup comparison of mean EMG activity during combined functional tasks. Each plot displays the EMG activity for a single muscle. Coloured horizontal bars indicate significant differences between groups (p < 0.05), while vertical bars represent the progression of the movement.

### 2.3. Data recording

The activity of 6 muscles per shoulder was recorded using a wireless EMG system (TRIGNO, Delsys Inc., USA) sampled at 2000 Hz (bandwidth 20–450 Hz, CMRR > 80 dB at 60 Hz, input impedance < 10 ohm, baseline noise < 750 nV RMS, effective EMG signal gain 909 V/V ± 5%). Surface EMG sensors were placed over the anterior (DELTA), middle (DELTM), and posterior (DELTP) deltoid, the upper (TRAPS) and middle (TRAPM) trapezius, and the serratus anterior (SERRA). Muscle selection was guided by superficial location, degree of implication in shoulder mobility and stability, and the technical limitation on the number of muscles that could be recorded with the available sensors. Skin preparation and sensor location followed the guidelines of the SENIAM [17]. To identify arm motion cycles, a set of reflective cutaneous markers were placed on the thorax, the arm, and the forearm (Fig. 1) following the recommendations of the International Society of Biomechanics (ISB) [50]. Their 3D trajectories were recorded using a 11-camera optoelectronic system sampled at 100 Hz (Miqus M3, Qualisys, Sweden).

### 2.4. Data processing

#### 2.4.1. Signal selection

A selection procedure was applied first to ensure data quality and comparability. EMG signals affected by motion artifacts were either excluded or corrected using an outlier removal method based on the standard deviation (SD) of the filtered signal (threshold set at 6 SD). Particular attention was also given to the cardiac interference on the left SERRA signal. To ensure methodological consistency across muscles, only signals with minimal contamination (<15% of normalised activity) were retained. The quality of the retained signals was assessed using the signal-to-noise ratio [43], with a minimum threshold of 10 dB set to ensure the validity of EMG analyses [23].

#### 2.4.2. Electromyographic signals

Retained raw signals were pre-processed following a standardised three-step procedure [53]: (1) fourth-order Butterworth band-pass filtering (15–475 Hz), (2) full-wave rectification, and (3) smoothing using a root mean square (RMS) with a moving window of 250 ms to obtain a stable and representative envelope of the muscle activation [39]. A signal normalisation was then performed by expressing each functional task signal as a percentage of the related maximum RMS value obtained during analytical tasks. This approach, corresponding to a submaximal normalisation, is particularly suitable for populations with functional limitations, as it offers a more functionally relevant alternative to isometric maximal voluntary contraction-based normalisation [3]. To detect arm motion cycles, thoracohumeral joint rotations were computed using a Y–X–Y Euler sequence decomposition, following the ISB recommendations [50]. The elevation angle was extracted, and a threshold-based method was applied to identify individual cycles. Finally, the processed EMG matrix **D** was stored. Due to clinical constraints and participant pain, the number of repetitions per movement was limited. To enhance motor variability, four distinct functional movements were selected, each performed three times, resulting in a total of 12 movement cycles. Consequently, the matrix **D** had dimensions 6L×L12,000, representing EMG signals from 6 muscles over 12 cycles of 1,000 time points each. All data processing was performed using MATLAB (R2024b, The MathWorks, USA).

#### 2.4.3. Muscle synergies

The muscle synergy analysis was performed using a non-negative matrix factorisation approach [25] implemented in MATLAB [20,35]. The processed EMG matrix **D** was factorised into the matrices **W** (muscle weightings) and **H** (temporal activations) following the relation **D** = **W** × **H**. Reconstruction quality was assessed using the variance accounted for (VAF), evaluated both globally (VAF_g) and individually for each muscle (VAF_m) [4,44]. Thresholds of 90% and 75% were used, respectively, to determine the optimal number of synergies that best explained the variance of the recorded signals. The algorithm was based on the approach proposed by Cherni et al. [5] and was implemented using the multiplicative update rules introduced by Lee & Seung [24]. To assess synergy stability, the extraction was repeated using a bootstrap procedure [6,42]. To obtain a global synergy profile, individual **W** matrices were grouped using k-means clustering. This approach allowed for the identification of dominant motor patterns within each group.

### 2.5. Statistical analysis

Preliminary analyses using the Shapiro–Wilk test confirmed that EMG data were normally distributed in both groups, whereas synergy data did not follow a normal distribution. Accordingly, parametric tests were applied to the EMG data, while non-parametric tests were used for the synergy-related analyses. Comparisons were performed using a one-way analysis of variance (ANOVA or Kruskal-Wallis test), followed by post-hoc tests (Student’s t-test or Wilcoxon/Mann-Whitney) where appropriate. Threshold of significance was set at 0.05. At the group level, mean muscle weightings were compared for each synergy, and mean muscle activations were analysed using one-dimensional Statistical Parametric Mapping (SPM1D), which enable the identification of specific time periods within the mean movement cycle where significant between-group differences occurred [32]. Additionally, the cosine similarity [19] was used to compare each individual symptomatic synergy profile to the mean asymptomatic synergy profile. Similarity scores range from 0, indicating no resemblance in curve shape, to 1, indicating identical profiles. A threshold of 0.8, commonly used in previous studies [31], was applied to determine whether the two synergy modules could be considered sufficiently similar. At the individual level, the cosine similarity was used to compare each symptomatic synergy profile to the mean asymptomatic synergy profile.

## 3. Results

### 3.1. Participants

Twenty patients (20 shoulders) were included in both the asymptomatic and symptomatic groups. The asymptomatic shoulders were not necessarily contralateral to the symptomatic ones. No significant differences were found between groups regarding age, body mass, height, or gender distribution (p < 0.05). All symptomatic shoulders received a rTSA using the Medacta Shoulder System. Group characteristics and surgical parameters are detailed in Table 1.

**Table 1.** Characteristics of the groups.

|  | Asymptomatic group<br>(n = 20) | Symptomatic group<br>(n = 20) | Assumption check |
| --- | --- | --- | --- |
| | Mean ± SD | Mean ± SD | p-value |
| Gender (female) | 12 | 10 | 0.541 |
| Age (yrs) | 71.7 ± 9.7 | 71.8 ± 10.3 | 0.975 |
| Body mass (kg) | 73.2 ± 14.8 | 77.4 ± 14.1 | 0.364 |
| Height (cm) | 164.5 ± 7.8 | 164.7 ± 7.6 | 0.919 |
| Constant-Murley score | - | 41.9 ± 10.5 | - |
| Humeral component inclination (°) | - | 132.2 ± 3.6 | - |
| Humeral component retrotorsion (°) | - | 32.7 ± 10.9 | - |
| Glenoid component inclination (°) | - | -4.4 ± 1.4 | - |
| Glenoid component retroversion (°) | - | 0.7 ± 1.4 | - |
| Glenosphere size (mm) | - | 37.7 ± 1.5 | - |

### 3.2. Electromyographic signals

The preoperative EMG analysis revealed a significantly higher muscle activity in the symptomatic group compared to the asymptomatic group (Fig. 1) for the DELTA ([0-13%] of the cycle, p = 0.007; [73-77%], p = 0.041; [95-100%], p = 0.046), DELTM ([0–100%], p < 0.001), TRAPS ([0-46%], p < 0.001; [53-56%], p = 0.047; [61-65%], p = 0.044; [78-100%], p = 0.002), and SERRA ([0–100%], p < 0.001). Postoperatively, some increases were observed for the DELTM ([34-56%], p = 0.003) and TRAPS ([29-36%], p = 0.039). Comparisons between pre- and postoperative measurements indicated a significantly higher preoperative muscle activity in the DELTM ([0-14%], p = 0.005; [60-62%], p = 0.048; [63-100%], p < 0.001), TRAPM ([0-17%], p = 0.032; [56-93%], p < 0.001; [96-100%], p = 0.049), TRAPS ([0-15%], p = 0.003; [93-100%], p= 0.039), and SERRA ([0-14%], p = 0.006; [79-100%], p = 0.002). The ratio between the TRAPS and SERRA was 1.7L±L1.4 in the asymptomatic shoulders, 1.6L±L1.7 preoperatively, and 1.4L±L1.0 postoperatively in the symptomatic shoulders. No significant differences were found across groups (ANOVA: F = 0.336, p = 0.716).

### 3.3. Muscle synergies

In the asymptomatic group, three motor clusters were identified and consolidated around the most representative ones. In contrast, the symptomatic group demonstrated a single dominant motor strategy, observed both pre- and postoperatively, making this clustering unnecessary. This analysis resulted in the extraction of two distinct muscle synergies (Fig. 2), each corresponding to a phase of arm movement common to both groups. The first synergy exhibited a peak of activity during the mid-elevation phase (around [20–40%] of the cycle), whereas the second synergy showed a biphasic pattern, with an increased activity at the beginning ([0–20%]) and end ([80–100%]) of the cycle, corresponding to the initiation and termination of the lowering phase. Despite similar temporal activation profiles, significant differences in muscle weighting were observed between groups. Preoperatively, the DELTP contribution in the first synergy was significantly higher in the symptomatic group compared to the asymptomatic group (pL<L0.001), while the TRAPS contribution was significantly lower (pL<L0.001). In the second synergy, this pattern was reversed: the DELTP showed a lower contribution (pL<L0.001), and the TRAPS a higher contribution in the symptomatic group (pL<L0.001). Postoperatively, no significant differences were observed for either synergy, although the DELTP contribution in the first synergy tended to be higher in the postoperative group, with a p-value close to significance (pL=L0.053). Comparisons between pre- and postoperative measurements indicated a significantly higher contribution of the DELTP in the first synergy preoperatively (p = 0.017), while the TRAPS was significantly lower (p < 0.001). In the second synergy, this pattern was reversed: the DELTP showed a lower contribution (pL<L0.001), and the TRAPS a higher contribution preoperatively (p < 0.001).

**Figure 2:**
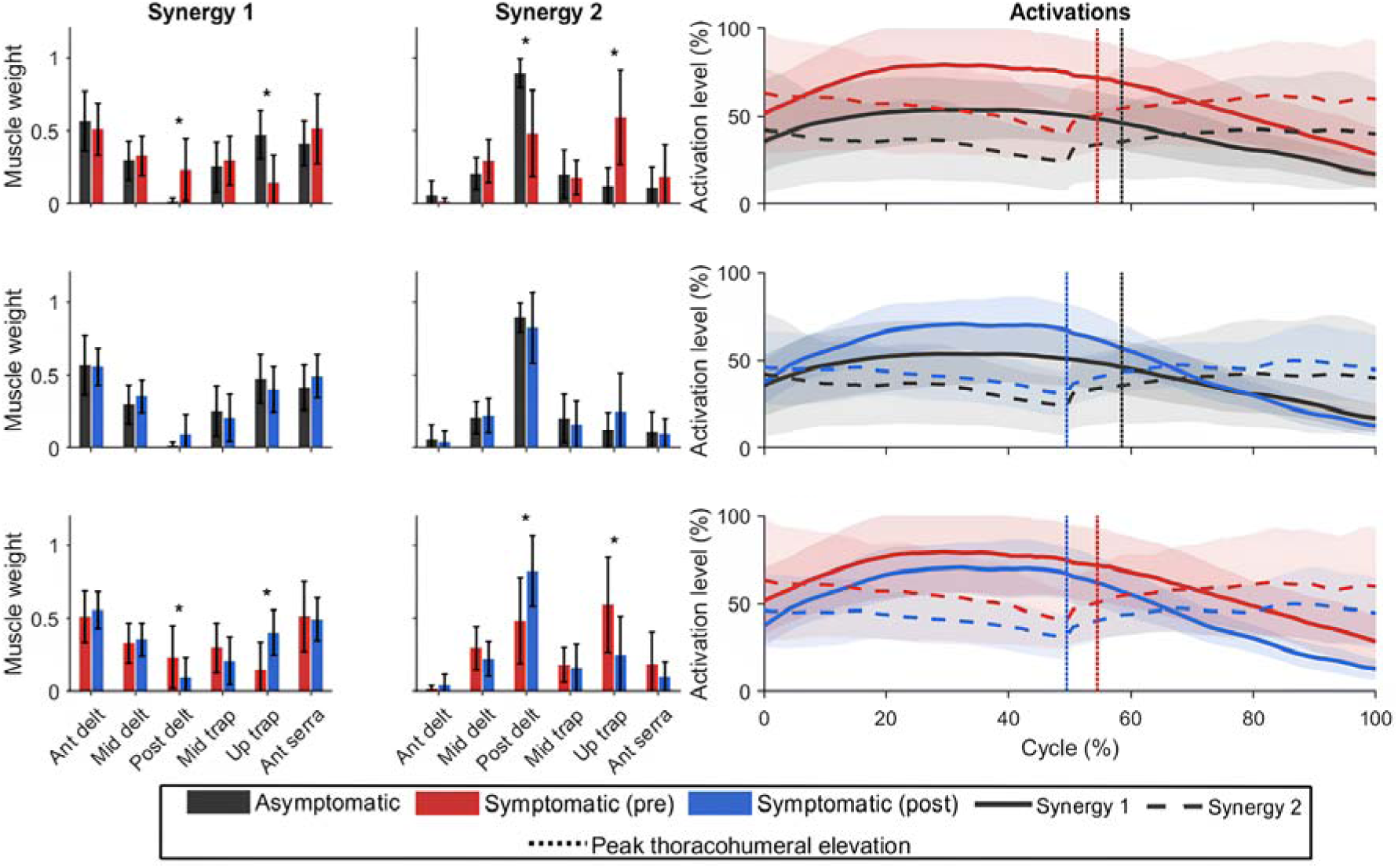
Intergroup comparison of mean muscle synergy profiles and temporal activations during combined functional tasks. Vertical bars represent the progression of the movement during the temporal activation of the synergies. DELTA: Anterior deltoid; DELTM: Middle deltoid; DELTP: Posterior deltoid; TRAPM: Middle trapezius; TRAPS: Upper trapezius; SERRA: Anterior serratus. *: p < 0.05.

A mean cosine similarity increase was observed between pre- and postoperative measurements (Fig. 3), with improvements of 15.96% for the first synergy and 37.42% for the second synergy after surgery. Postoperatively, 17 symptomatic shoulders (85%) for synergy 1 and 15 shoulders (75%) for synergy 2 demonstrated muscle activation profiles more similar to those of asymptomatic shoulders, while the remaining shoulders showed no improvement or a decrease. Significant differences in the mean cosine similarity were found between the preoperative symptomatic and asymptomatic groups (pL<L0.001), as well as between pre- and postoperative measurements within the symptomatic group (pL<L0.001). In contrast, no significant differences were observed between the postoperative symptomatic and asymptomatic groups (pL=L0.818 for synergy 1, and pL=L0.457 for synergy 2).

**Figure 3:**
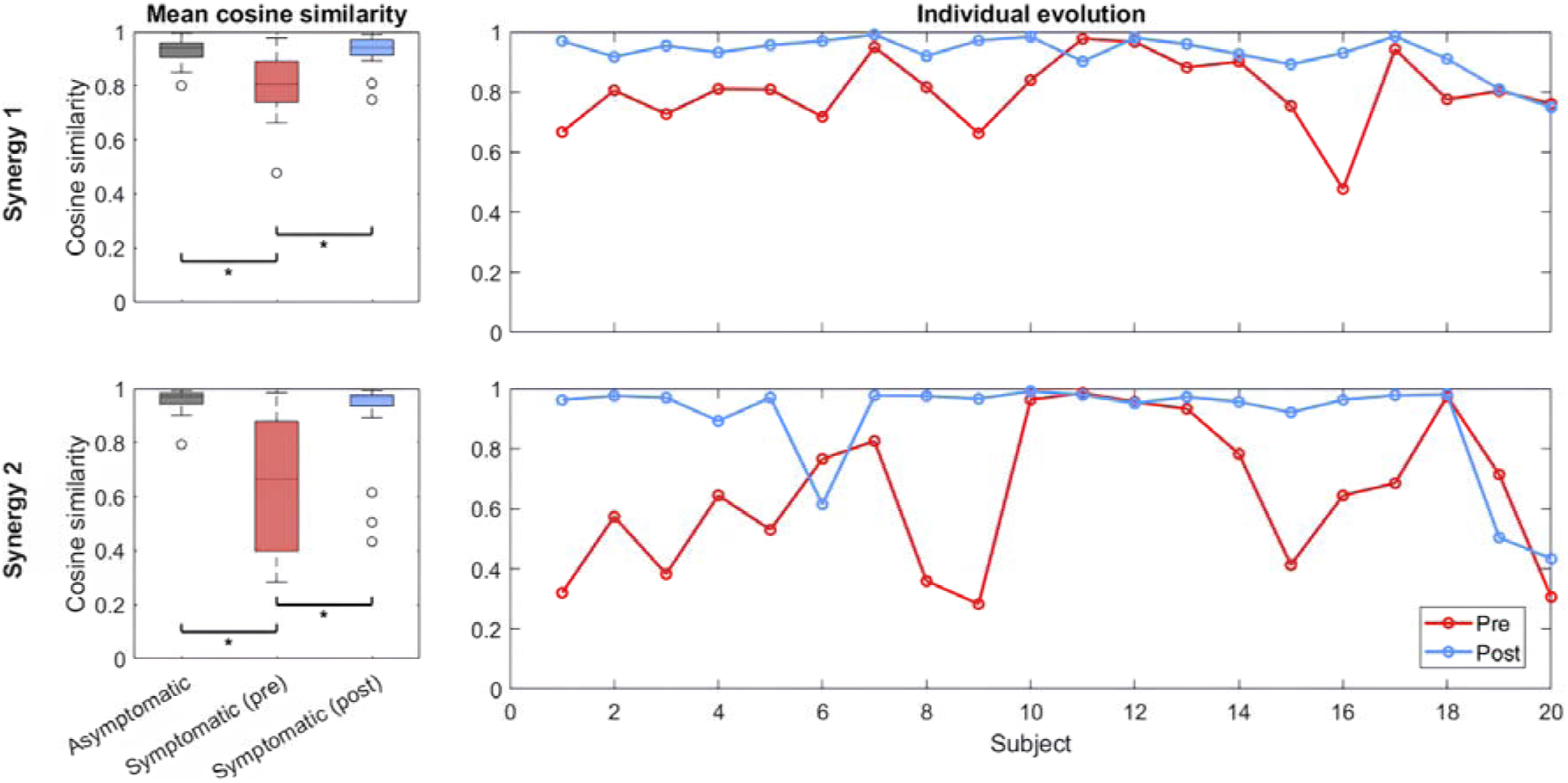
Comparison of individual preoperative and postoperative synergy profiles with the asymptomatic mean profile during combined functional tasks. o: outliers and *: p < 0.05.

## 4. Discussion

This study investigated changes in superficial shoulder muscle activity and synergy following rTSA, using asymptomatic shoulders as a reference. Preoperatively, participants with symptomatic MRCT showed an increased muscle activation, particularly in the DELTM, TRAPS, and SERRA. Muscle synergy patterns also differed from the asymptomatic group, with altered DELTP weightings during arm elevation and increased TRAPS activation during lowering, as evidenced by reduced similarity scores. Postoperatively, muscle activity patterns are more closely aligned with those of the asymptomatic group, although the TRAPS and DELTM activation remained abnormally high. Synergy patterns also showed a greater similarity to the asymptomatic group. These findings support the hypothesis that rTSA does not significantly alter the number or structure of muscle synergies but facilitates a reorganisation of coordination patterns toward asymptomatic profiles at both group and individual levels.

### 4.1. Stability of the number of synergies

The number of extracted synergies did not differ significantly among asymptomatic, MRCT, and rTSA shoulders. Although often considered as a global indicator of motor control complexity [7], this parameter alone may not fully capture the extent of underlying neuromuscular adaptations. Some studies have reported an increased number of synergies in pathological shoulders [14], suggesting a disruption in motor control strategies. In contrast, other studies [53] have found no significant differences between symptomatic and asymptomatic groups, implying that the number of synergies may remain stable despite musculoskeletal impairments. This suggests that the number of synergies is a relatively insensitive metric, which might explain its limited responsiveness to rehabilitation. Similarly, even following surgery, as in our study, the number of synergies remained stable between MRCT and rTSA shoulders. Nonetheless, surgery appears to induce a reorganisation of the neuromuscular coordination strategies, as evidenced by the significant changes observed in both muscle weightings and EMG activation patterns, particularly in the scapular and deltoid muscles.

### 4.2. Scapular and deltoid adaptations reflecting neuromuscular coordination strategies in massive rotator cuff tears

In symptomatic MRCT, our results revealed significant changes in the TRAPS, SERRA, and DELTP, suggesting major adaptations in muscle recruitment strategies. Supporting this, the systematic reviews by de Oliveira et al. [11], and Spall et al. [40], reported substantial alterations in the shoulder muscle activity in individuals with rotator cuff deficiencies, particularly an increased recruitment of the superficial shoulder muscles to compensate for an impaired cuff function. Specifically, we observed an increased and prolonged activation of the TRAPS, consistent with previous findings [12,16,40]. This hyperactivity is often associated with muscle pain, cervical tension, and headaches [29]. The SERRA adaptations observed preoperatively also reflect a shift in the functional role of scapular stabilisers, as described in prior studies [12,16]. Such neuromuscular changes may contribute to scapular dyskinesis [37,41], even though no significant between-group differences were observed in our dataset concerning the ratio of activity between the TRAPS and SERRA. Our findings also align with the compensatory mechanisms described by Spall et al. [40], who reported both a redistribution of muscle activity toward scapular stabilisers and an increased co-contraction of adductors to enhance the glenohumeral stability in symptomatic patients. The TRAPS was no longer predominantly involved during the first synergy related to arm elevation but was instead mainly active during the second synergy related to arm lowering. This change may primarily result from the reduced arm elevation observed preoperatively, which prevents the activation of this muscle typically recruited during the final portion of elevation [9,12]. Consequently, the TRAPS appears to be redirected toward controlling arm descent and scapular stabilisation. Concerning the DELTP, although our results did not reveal significant changes in terms of muscle activation pattern, the synergy analysis highlighted a higher contribution of this muscle to the synergy related to arm elevation. According to Veen et al. [47], this may reflect a subtle compensatory mechanism, characterised by DELTP overactivation and a disrupted coordination with the other deltoid heads during functional tasks, aiming to support arm elevation despite rotator cuff deficiencies. These findings align with previous studies [11,47] and support the idea that neuromuscular adaptations can occur without being detectable through the analysis of raw EMG signal amplitudes alone. Instead, they emerge through changes in muscle coordination patterns, which are more effectively revealed by the synergy analysis.

### 4.3. Deltoid adaptations and inter-individual variability reflecting neuromuscular coordination strategies after reverse total shoulder arthroplasty

Following rTSA surgery, our results revealed a functional reorganisation consistent with the biomechanical objectives. Notably, the DELTM increased activation reflects its enhanced role in arm elevation. This observation is consistent with the findings of Pietroski et al. [33], who reported a sustained increase in the DELTM activation six months after rTSA, particularly during abduction and flexion, reinforcing its compensatory role in generating elevation joint moment and stabilising the humeral head in the absence of a supraspinatus function. This adaptation is even facilitated by the lateralised prosthesis design used, which displaces the glenohumeral joint centre and enhances the deltoid’s lever arm, thereby enabling more effective joint moment production [21]. These functional improvements, also reflected in synergy profiles, may help mitigate the TRAPS compensatory overactivation and contribute to the postoperative pain reduction [30]. In both synergies, muscle weightings shift toward the asymptomatic profiles, suggesting a beneficial effect of the surgery in restoring a physiological motor control [52]. While synergies tended to approach asymptomatic profiles, inter-individual variability persisted, especially in the DELTP, which showed a higher contribution for the first synergy in several cases. This trend, although not statistically significant, may reflect an attempt at a compensatory reorganisation aimed at restoring a balanced deltoid recruitment [38]. Although most participants showed an increased cosine similarity with the asymptomatic profile, some of them continued to display lower scores. These differences highlight the influence of inter-individual variability on postoperative neuromuscular recovery. While surgical factors such as the implant design and humeral inclination can affect muscle recruitment [8,13,18], all patients in this study underwent a standardised implant positioning. Additionally, the age and preoperative status may influence recovery dynamics [22,48], but do not sufficiently account for the variability observed. Some neurological pathologies, particularly the arthrogenic muscular inhibition, may also play a key role [26]. Indeed, this pathology, triggered by altered joint afferents, leads to a reflexive and central inhibition of periarticular muscles, potentially impairing the voluntary activation. This persistent inhibition may then prevent the full voluntary recruitment of periarticular muscles such as the deltoid and scapular stabilisers, despite an optimal implant positioning and successful structural outcomes. All these factors are likely to contribute to the observed heterogeneity and underscore the need for similar studies in larger samples to validate these findings.

### 4.4. Limits of the study

This study has some limitations. First, the synergy analysis relies on a set of motor tasks to capture coordination patterns, but the limited number of movement repetitions analysed may have influenced the results. However, in preoperative shoulders, pain and/or restricted mobility sometimes prevented task execution, thereby limiting completion of further movement cycles. Second, although the selected tasks were functional, they did not fully reflect the complexity of everyday shoulder movements. To address this limitation, we deliberately combined several functional tasks and prioritised those involving higher levels of intermuscular coordination. Third, the potential influence of the postoperative rehabilitation on motor control adaptations was not analysed. Although therapeutic approaches were generally similar across patients, rehabilitation is known to impact neuromuscular coordination [49]. Future studies are warranted to better distinguish the respective contributions of surgery and rehabilitation to the reorganisation of muscle coordination.

## 5. Conclusion

Synergy analysis highlights neuromuscular adaptations in symptomatic massive rotator cuff tear planned for reverse total shoulder arthroplasty with a reorganisation involving the posterior deltoid and upper trapezius, reflecting a return toward a functional motor pattern. The greater involvement of the middle deltoid aligns with the surgical goal of rebalancing shoulder function, although this pattern was not reflected in the synergy analysis. The observed inter-individual variability in patients’ recovery raises the need to better consider clinical profiles and rehabilitation.

## 5.1. Data availability

Data are available upon request to the corresponding author.

## 5.4. Authors contributions

**HF**: Conceptualisation, Formal analysis, Methodology, Visualisation, Writing – original draft. **NH**: Conceptualisation, Manuscript revision. **FM**: Conceptualisation, Data collection, Methodology, Visualisation, Supervision, Manuscript major revision. **YC**: Conceptualisation, Methodology, Software, Visualisation, Supervision, Manuscript major revision.

